# Can Dental AI Really Beat Dentists? DentalPair-Cert for Rigorous AI–Dentist Inference

**DOI:** 10.64898/2026.09.01.26361874

**Authors:** Shahran Rahman Alve, Sunehera Rahman, S M Abeer Chowdhury Meem

## Abstract

A dental AI system and a dentist reading the same radiographs form a paired comparison. Published comparative studies often report the two arms separately against a reference standard, leaving the joint pattern of correctness between them unavailable for secondary paired inference. We show what that omission costs. The accuracy difference remains exactly identified; its sampling variance does not, so the report contains the estimate and not its uncertainty. On a study of 282 units, two published accuracies are consistent with 38 distinct joint tables whose confidence intervals differ in width by a factor of 2.5. The consequence is a three-zone decision map rather than a single threshold: differences at or below 1.06 points are non-significant under every compatible table, differences at or above 6.03 points are significant under every compatible table, and in between the published numbers cannot decide. We then show the omission is repairable at negligible cost. One additional integer, the number of units both arms classify correctly, identifies the joint table exactly and restores standard paired inference. For a panel of readers the pairwise dependences must arise from one joint distribution, a constraint that binds once three readers are present; publishing each reader’s joint-correct count against a single reference reader cannot widen and may tighten every pairwise bound, and in a 7-arm experiment reduced them by a median of 37% even for pairs excluding that reference. Where the integer was never published we give DentalPair-Cert, an interval with finite-sample coverage uniformly over every admissible within-unit AI–dentist dependence under the independent-sampling-unit model, certified in both the nuisance maximisation and the inversion. Across 4,200,000 simulated comparisons an independence analysis falls to 74.5% coverage with 12.2% type-I error; in a purposive sample of 9 recent comparative studies, 1 reported a paired test on discordant units.

## I. Introduction

Papers claiming that an AI system reads dental radiographs as well as, or better than, a dentist now appear at a steady rate. The claim rests on a comparison, and the comparison rests on a design: the same images are shown to both arms, and both are scored against a reference standard. Reading the same images is what makes such a study efficient. Two readers facing an identical set of difficult cases share that difficulty, and the shared part cancels when their accuracies are compared.

The cancellation is invisible in the published tables. A typical report gives the AI system’s sensitivity, the dentist’s sensitivity, the denominators, and a p-value against the reference standard for each arm separately. What it omits is how often the two arms were right together, wrong together, or split. Write the joint table of correctness as *a* units both arms classify correctly, *b* correct for the AI alone, *c* correct for the dentist alone, and *d* neither. The published marginals fix *x*_1_ = *a* + *b* and *x*_2_ = *a* + *c*. They say nothing about how *x*_1_ divides between *a* and *b*, and that division is what governs how precisely the difference between the arms is measured.

Two consequences follow, and they pull in opposite directions. Since *b* − *c* = *x*_1_ − *x*_2_ holds for every table compatible with the marginals, the accuracy difference is identified exactly. Since *b* + *c* = *x*_1_ + *x*_2_ − 2*a* is free, its sampling variance is not identified at all. A published pair of accuracies therefore reports a point estimate and withholds the uncertainty attached to it. An analyst who proceeds as though the two arms were independent has assumed a specific dependence, one the design neither produces nor makes checkable.

At the sample sizes dental reader studies actually use, the effect is large. Consider a study of 282 units in which the AI is correct on 245 and the dentist on 231. Exactly 38 joint tables are consistent with those numbers. The confidence interval a reader could compute ranges from 0.0518 to 0.1309 in width across them, a factor of 2.5.

What follows is not a single threshold but three zones (Section III-C). A sufficiently small difference is non-significant under every compatible table, so it is decided, and decided against significance. A sufficiently large one is significant under every compatible table. Between the two boundaries the published numbers are genuinely ambiguous, because some compatible tables imply significance and others do not. On a study of 282 units those boundaries sit at 1.06 and 6.03 accuracy points. To see that the middle zone is not a remote corner of the parameter space, we re-expressed the AI-minus-dentist differences reported by one audited study at that size: of 3 reader strata, 2 fall inside it (Section VIII).

None of this is a criticism of any individual study. It is a property of a reporting convention, and the convention has a cheap fix.

### A. Contributions

#### We quantify what a published pair of accuracies determines

The compatible tables form an integer interval with a closed-form length, and we compute the resulting spread of intervals across the sample sizes dental reader studies occupy (Section III).

#### We show one integer is sufficient

Any one of the four cell counts, or equivalently the agreement rate between the arms, identifies the whole table and restores exact paired inference. Nothing is estimated and no assumption is introduced (Section IV).

#### We extend the argument to reader panels

With *R* human readers plus one AI system, each sampling unit produces a correctness vector in {0, 1}^*R*+1^, and all pairwise dependences must be marginals of one joint distribution over its 2^*R*+1^ cells. We prove this restricts the admissible set once three arms are present, give a linear program for the sharp bounds, and show that publishing each reader’s joint-correct count against one reference reader cannot widen and may tighten every pairwise bound, including those not involving the reference (Section V).

#### We give a certified interval for studies already published

Where the integer is absent and cannot be recovered, DentalPair-Cert returns an interval with finite-sample coverage uniformly over every admissible within-unit AI–dentist dependence, under a model in which sampling units are independent. Both numerical steps, the maximisation over the unidentified nuisance and the inversion over the effect size, are computed by rigorous outer bounds, so the implementation inherits the guarantee rather than approximating it (Section VI).

#### We validate and then apply the analysis

Evidence is kept in three separate layers: a proved coverage guarantee, exhaustive finite-sample coverage computed at small *N*, and Monte Carlo operating characteristics over 4,200,000 replicates for the methods actually simulated. A controlled non-clinical benchmark, where the joint table is available, supplies real dependence structure (Section VII). We then examine 9 recent comparative studies and report what each makes recoverable (Section VIII).

## II. Related work

### A. Comparative dental AI studies

Recent work comparing dental AI against clinicians spans caries on bitewings [3], [5], periodontal bone loss [1], periapical and restorative findings [2], and panoramic images [4], [6], [7], [9]. The designs vary. Li *et al*. [1] used a randomised crossed reader design with a two-week washout and compared clinician against model with McNemar’s test on discordant pairs; among the studies we examined it is the only one reporting the comparison in that form. Kazimierczak *et al*. [8] report a head-to-head AI-versus-reader arm with bootstrap intervals, and Kwiatek *et al*. [5] publish AI-clinician concordance rates, which Section IV shows is an equivalent sufficient statistic. Others report per-arm accuracies against a consensus reference [3], [7] or a paired AUC test without the underlying counts [4]; Section VIII gives the tally. Shared corpora such as PRAD [17] make the reporting question more pressing, not less: once several systems are scored on one corpus, every comparison inherits the structure analysed here.

### B. Appraisals of the dental AI literature

Reviews of this literature have converged on reporting quality as the binding problem. Khubrani *et al*. [10] scored 30 studies with APPRAISE-AI and rated none very high quality; Saikia *et al*. [14] appraised 116 systematic reviews with ROBIS and restricted pooling to the twelve judged low risk of bias; Alaqla *et al*. [12] found bias concerns in 28 studies concentrated in the reference standard; and Hung *et al*. [15] applied CLAIM-adapted criteria to 43 studies and reported high risk on code accessibility in most. Albano *et al*. [11] and Luke and Rezallah [13] abandoned or qualified meta-analysis because of heterogeneity, and Shi *et al*. [16] note that few studies of jaw lesions compare against clinicians at all. These appraisals identify the gap; we supply the mechanism, the size of the loss, and a fix.

### C. Reporting guidance and paired inference

Guidance for AI diagnostic studies is well developed: STARD-AI [20], the CLAIM update [21] and REFORMS [22] all ask for comparative analyses respecting the study design, and Metrics Reloaded has shown such practice can be reformed by community guidance [23], [24]. Guidance governs future papers. It does nothing for those already published, which are the corpus a meta-analyst or guideline panel must work with now.

The statistical tools for paired binary comparison are also settled. McNemar’s test [25] and the interval methods of Newcombe [26], Tango [27] and Agresti and Min [28] are reviewed and benchmarked by Fagerland *et al*. [29], with recent refinements by Chang *et al*. [30] and Roldán-Nofuentes *et al*. [31]. Every one of them needs the discordant counts *b* and *c*. Multi-reader multi-case analysis [32], [33] solves a harder problem when reader-level data are available. We use these as measures of what full information buys, not as alternatives, since none can be computed from (*x*_1_, *x*_2_, *N*).

### D. Partial identification

When marginals are known and the joint is not, the Fréchet inequalities [34] bound the admissible joints and Manski [35] gives the general account of what such bounds support; Salako [36] develops constructive proofs of the generalised form. The structure recurs in ecological inference [37] and in meta-analysis when within-study correlations go unreported [38]. Papadopoulos and Fiska [39] derived Fréchet bounds for unobserved concordance in paired binary meta-analysis. Their target differs from ours: they bound a joint cell probability and carry an interval-valued estimand forward, whereas here the estimand is already point-identified and the sampling variance is what is lost. What must be built is therefore not an interval-valued parameter but a test valid uniformly over the unidentified nuisance, with an inversion that provably encloses the accepted set. Outside dentistry, the machine learning community has measured how strongly model errors correlate on shared test sets [40], [41] and added uncertainty to benchmark comparisons [42], [43], where per-item outputs exist. We treat the case where they do not.

## III. What a published pair determines

### A. Setup

Let *N* units be read by both arms, with *X*_*i*_, *Y*_*i*_ ∈ {0, 1} recording whether the AI and the dentist are correct on unit *i*. Write the cell probabilities (*π*_11_, *π*_10_, *π*_01_, *π*_00_), marginals *p*_1_ = *π*_11_ + *π*_10_ and *p*_2_ = *π*_11_ + *π*_01_, and the estimand Δ = *p*_1_ − *p*_2_. Under multinomial sampling,

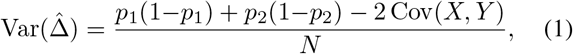

and with *q* = *π*_11_ the covariance is *q* − *p*_1_*p*_2_. The Fréchet inequalities max(0, *p*_1_+*p*_2_ − 1) ≤ *q* ≤ min(*p*_1_, *p*_2_) bound the unidentified term and give sharp bounds on the variance,

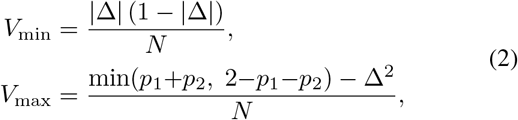

both attained by admissible joint distributions. The independence variance *V*_ind_ = [*p*_1_(1−*p*_1_) + *p*_2_(1−*p*_2_)]*/N* satisfies

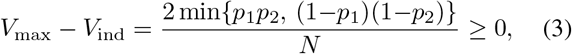

so independence sits strictly inside the admissible range. Assuming it is a choice about the dependence, not a conservative default.

### B. The compatible tables

At the level of counts the structure is exact and elementary. Every cell is an affine function of the single unknown *a*:

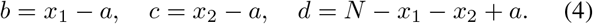

Non-negativity of the four cells confines *a* to the Fréchet range, so the number of compatible tables is

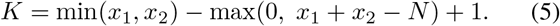

Over those *K* tables the discordance *b* + *c* sweeps its full range while *b* − *c* never moves. Fig. 1 shows the consequence for one study size: the point estimate is a horizontal line and the interval around it expands or contracts by a factor of 2.5 depending on a number the paper did not print.

**Fig 1.**
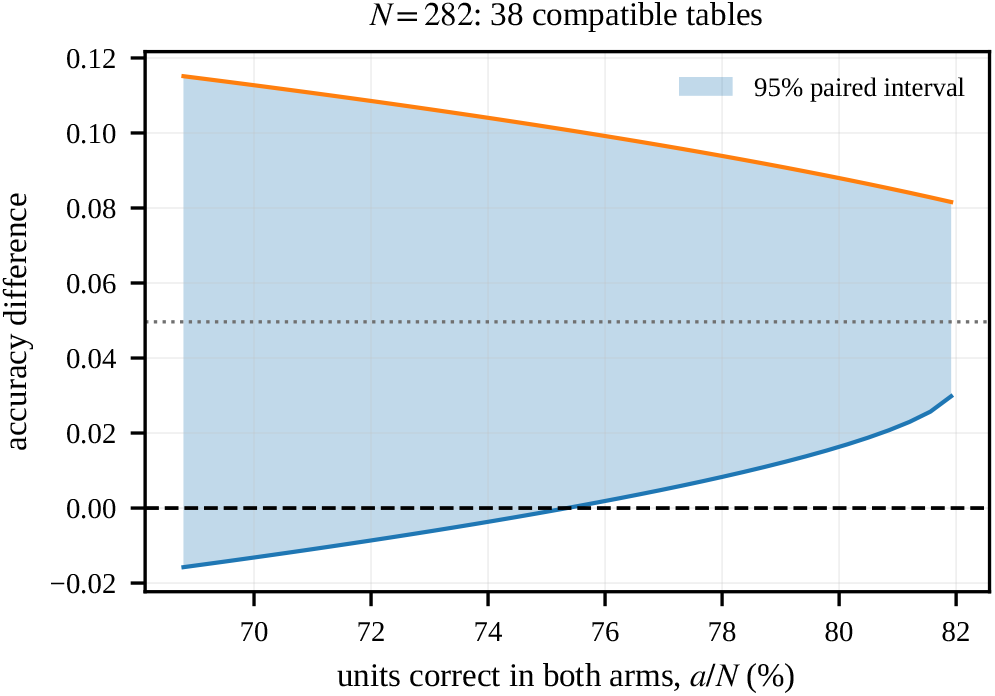
Every point on the horizontal axis is a joint table compatible with the same two published accuracies on a study of 282 units. The shaded band is the two-sided 95% Tango score interval, located by bisection that table implies; the dotted line is the point estimate, which does not move. Interval width varies by a factor of 2.5 across tables a reader cannot distinguish.

### C. Three decision zones

Throughout this subsection, significance and the compatible-table confidence intervals are defined by the two-sided 95% Tango score interval, located by bisection [27], and every value in Table I and Fig. 2 is generated by that same procedure. The choice matters: the numerical boundaries move if significance is instead defined by inverting McNemar’s test or by a Newcombe or Wald interval, because those procedures do not agree at a given table. We use the score interval because it was the best calibrated of the paired methods on our evaluation grid (Table IV). The implementation locates each endpoint by bisection rather than scanning a grid of candidate effects, since at these sample sizes a scan coarse enough to be affordable resolves the interval with only a few points; it is checked against the grid implementation wherever the grid is fine enough for the two to be compared.

**TABLE I.**
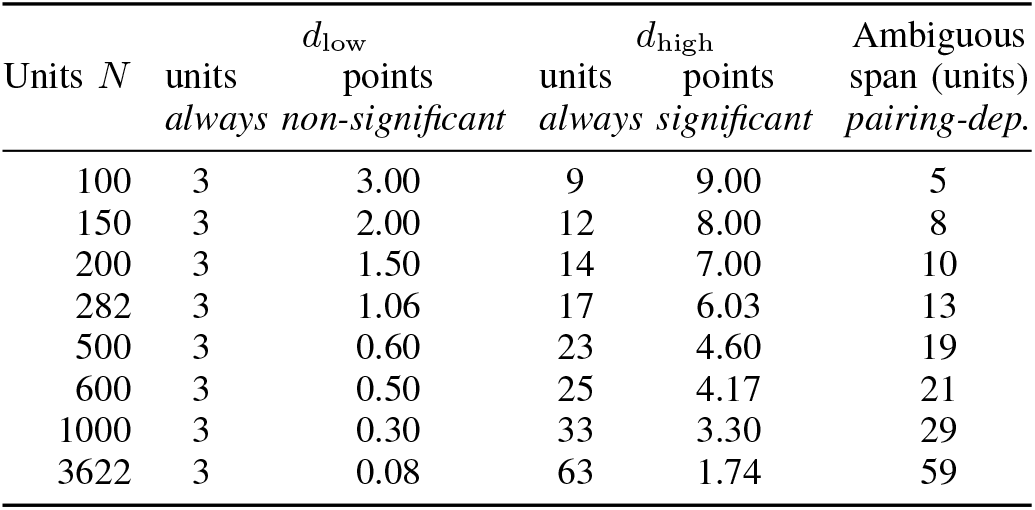
the three decision zones at 85% baseline accuracy, all computed with the two-sided 95% tango score interval, located by bisection. at or below *d*_low_ the comparison is *always non-significant*: every compatible joint table agrees. At or above *D*_high_ it is *always significant*. Between them it is *pairing-dependent*, and the published marginals cannot decide. Points are percentage points of accuracy.

**Fig 2.**
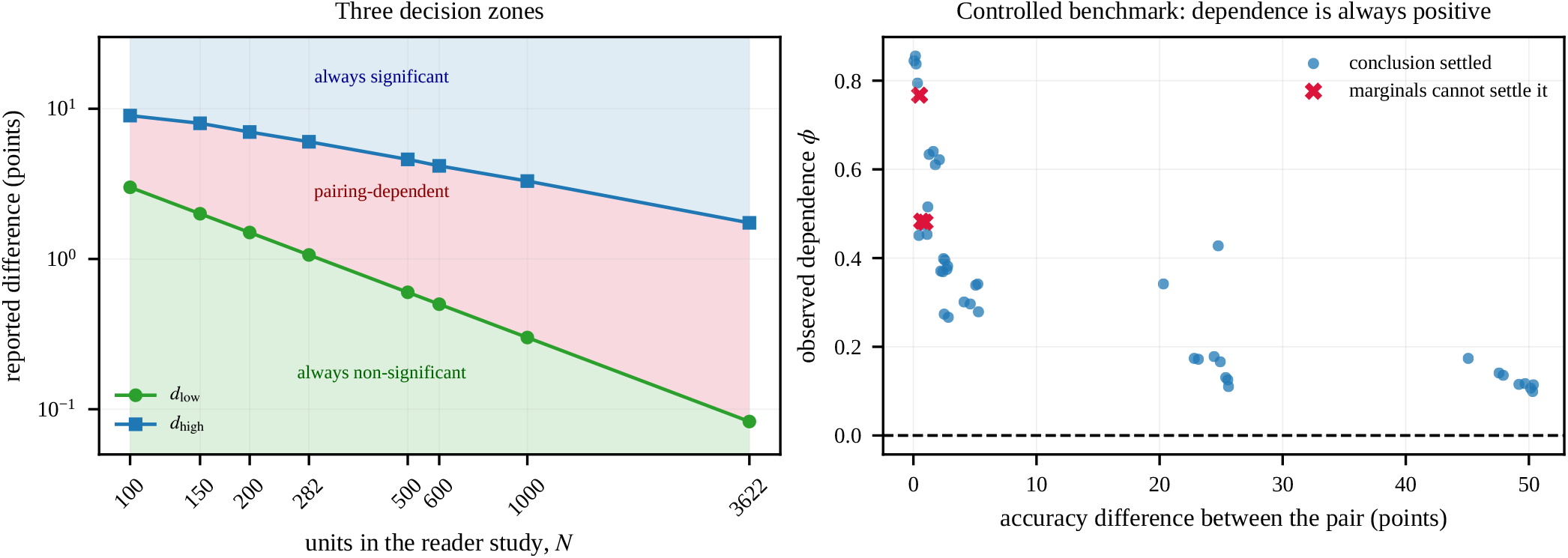
Left: the three decision zones against study size, at 85% baseline accuracy, computed with the two-sided 95% Tango score interval, located by bisection and shaded: at or below *d*_low_ every compatible joint table is non-significant, at or above *d*_high_ every one is significant, and between them the published numbers cannot decide. Right: dependence measured between 45 pairs of classifiers scored on the same 1,797 non-clinical images. It is positive in every pair, and the comparisons the marginals cannot settle are the close, strongly dependent ones.

The width of the paired interval increases with the discordance *b* + *c* = *x*_1_ + *x*_2_ − 2*a*, so across the compatible tables it is widest at *a* = *a*_min_ and narrowest at *a* = *a*_max_. Examining those two endpoints is therefore exact rather than a heuristic, and it yields two boundaries rather than one.

Write *d* = *x*_1_ − *x*_2_ for the reported difference. Let *d*_low_ be the largest *d* at which even the *narrowest* compatible interval still contains zero, and *d*_high_ the smallest *d* at which even the *widest* compatible interval excludes it. Then:

- *d* ≤ *d*_low_: every compatible table is non-significant. The published numbers do settle the question, against significance.
- *d*_low_ *< d < d*_high_: some compatible tables imply significance and others do not, so the published numbers cannot decide.
- *d* ≥ *d*_high_: every compatible table is significant.

An earlier form of this analysis reported only *d*_high_ and described everything below it as ambiguous. That is one-sided: it wrongly places arbitrarily small differences in the ambiguous zone when they are in fact decided. Both boundaries are located by exact scan over *d*, and the scan verifies that the two significance indicators are monotone in *d* and mutually consistent. The exact scan confirmed monotonicity and consistency of the two significance indicators in every case evaluated, so the boundaries are computed rather than assumed.

Table I gives both boundaries across the range of sizes dental reader studies use, and Fig. 2 (left) plots the three zones. Two features are worth noting. The upper boundary falls with *N* as one would expect, from 9.00 points at *N* = 100 to 1.74 points at *N* = 3622. The lower boundary is a fixed *count* rather than a fixed rate: it sits at 3 units at every size we examined, because at maximum concordance all discordance is one-directional, so the narrowest compatible interval depends on the difference through a count of discordant units and not through *N*. The ambiguous zone therefore widens in absolute terms as studies grow, from 5 units at *N* = 100 to 59 units at *N* = 3622, even as it narrows in percentage points.

## IV. One integer is sufficient

### Theorem 1

(One additional count is sufficient). *Given x*_1_, *x*_2_ *and N, each of a, b, c, d determines the other three. So does the total agreement rate* (*a*+*d*)*/N, which counts both the both-correct and both-wrong states, and so does the discordance b* + *c. Publishing any one of them identifies the joint table exactly, and with it* 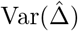 *and every standard paired test and interval*.

*Proof*. By (4) the map *a* ⟼ (*a, x*_1_ − *a, x*_2_ − *a, N* − *x*_1_ − *x*_2_+*a*) is a bijection from the admissible range onto the tables with the given marginals, and *b, c, d* are invertible affine functions of *a*. For the agreement rate, *a* + *d* = 2*a* + *N* − *x*_1_ − *x*_2_, so

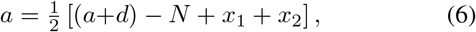

which recovers *a* from the agreement count. Since *b* + *c* = *N* − (*a* + *d*), the discordance determines it as well.

□

The algebra is elementary. What matters is the practical reading. The information missing from a comparative dental AI study is exactly one integer. Obtaining it requires no additional imaging, no further reading session and no re-consent: the number already exists inside any analysis that scored two arms on the same units, and is discarded when the results are written up. Reporting it converts a comparison whose conventional paired uncertainty is unidentified into one that supports the exact paired analysis the field already knows how to perform [29].

We recommend *a*, the count correct in both arms, because it is the most directly interpretable of the four and is bounded above by the smaller of the two accuracies. The agreement rate is equally sufficient by (6), and at least one recent study already reports it [5], which shows the recommendation asks for nothing unusual.

## V. Reader panels

Dental reader studies rarely use one dentist. A panel of *R* readers plus an AI system produces, on each unit, a correctness vector in {0, 1} ^*R*+1^, and all the pairwise dependences must be marginals of one joint distribution over its 2^*R*+1^ cells. Treating each AI-versus-reader comparison in isolation is valid but discards that constraint, which is real.

### Proposition 1

(Pairwise extremes need not be jointly attainable). *For three or more arms there exist marginals such that each pairwise dependence can individually reach its Fréchet extreme, while no joint distribution reaches all of them at once*.

*Proof*. Take three arms with *p*_1_ = *p*_2_ = *p*_3_ = 1*/*2. Each pair can be fully discordant on its own. Suppose all three were simultaneously. Full discordance of arms 1 and 2 forces *X*_2_ = 1 − *X*_1_ on every unit, and of arms 1 and 3 forces *X*_3_ = 1 − *X*_1_. Then *X*_2_ = *X*_3_ everywhere, so arms 2 and 3 are fully concordant, contradicting full discordance of that pair.

□

We confirm Proposition 1 numerically as well as on paper. The linear program below, asked for a joint distribution realising all three extremes together, reports the system infeasible.

### A. A linear program over the joint

Every quantity we need is a linear functional of the joint, so the sharp bounds come from linear programming over the 2 ^*R*+1^ simplex. With variables *p*_*S*_ ≥ 0 for *S* ⊆ {1, …, *R*+1} and∑_*S*_*p*_*S*_ = 1, impose interval constraints on each marginal, *l*_*i*_ ≤ ∑_*S*∋*i*_ *p*_*S*_ ≤ *u*_*i*_, and, where reported, on pairwise joint-correct probability, *l*_*ij*_ ≤ ∑ _*S⊇*{*i,j*}_ *p*_*S*_ ≤ *u*_*ij*_. That sum is *P* (*X*_*i*_=1, *X*_*j*_=1), the probability both arms are correct. It is not total agreement *P* (*X*_*i*_=*X*_*j*_), which would also include the both-wrong state; the two are interconvertible given exact marginals but not under marginal confidence boxes, so the program, the published statistic and the results below all use the joint-correct count. Maximising ∑_*S*: |*S*∩{*i,j*}|=1_ *p*_*S*_ returns a sharp upper bound on the discordance probability of the pair (*i, j*), which is the quantity that caps the nuisance in Section VI.

Two details make this usable. The published accuracies are estimates, so we pass a simultaneous Bonferroni–Clopper– Pearson box for the marginals into the program as interval constraints; one solve then bounds the discordance over the whole box, with no search over it. And because the constraints couple the pairs, partial reporting propagates. As a check on the formulation, when only marginals are supplied the program reproduced the analytic single-pair Fréchet cap exactly, as it must.

### B. One reference reader

This suggests a protocol that scales with the panel. Rather than an integer for each of the 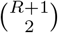 pairs, publish each reader’s joint-correct count against one designated reference reader: *R* integers for the whole panel. Adding constraints to a maximisation cannot enlarge its feasible set, so at theorem level the statement is that every pairwise bound *cannot widen and may tighten*, including bounds between two readers neither of which is the reference, which are constrained only through transitivity. Whether the tightening is strict for a given pair is an empirical matter.

Section VII measures this on joint-correct counts that actually occurred. In that 7-arm experiment every evaluated bound tightened strictly, and 6 additional integers reduced the certified discordance cap by a median of 57% for pairs involving the reference and 37% for pairs that do not, the latter reaching 51%. Those are results from this experiment, not general guarantees. Which arm serves as reference matters: across the 7 possible choices the median reduction for non-reference pairs runs from 7% to 37%. A central, high-accuracy reference is worth substantially more than a peripheral one (Table II), and the choice is made once, before any reporting.

**TABLE II.** Reduction in the certified discordance cap from publishing one reference *joint-correct* column (6 integers for the 7-arm panel), by choice of reference. The statistic is the number of units both arms classify correctly, *P* (*X*_*i*_=1, *X*_*j*_ =1), and not total agreement. Joint-correct counts are those measured in Section VII.

| Reference arm | With ref. median (%) | Without reference median (%) | max (%) |
| --- | --- | --- | --- |
| knn_k5 | 57 | 37 | 51 |
| knn_k3 | 56 | 36 | 51 |
| knn | 56 | 35 | 49 |
| extra_trees | 57 | 33 | 40 |
| extra_trees_b | 56 | 30 | 38 |
| random_forest | 53 | 11 | 35 |
| random_forest_b | 52 | 7 | 33 |

## VI. DentalPair-Cert

For studies already published the integer is absent and no protocol recovers it retrospectively. What remains available is an interval valid uniformly over every admissible within-unit AI–dentist dependence, under the independent-sampling-unit model of Section III. Dependence between sampling units is a separate layer that this guarantee does not cover.

Bounds on a variance are not a confidence statement, so we work with the test. The structural fact we exploit is that the distribution of *T* = *x*_1_ − *x*_2_ = *b* − *c* depends on the four cell probabilities only through (*π*_10_, *π*_01_), since (*b, c, N* −*b*−*c*) is multinomial with those probabilities. Under *H*_0_ : Δ = Δ_0_ the nuisance collapses to the scalar *v* = *π*_01_, with *π*_10_ = *v* + Δ_0_. Maximising the exact two-sided p-value of *T* over *v* and inverting yields a set valid for any dependence, because the true *v* is among the values maximised over.

Maximising over the whole feasible range is valid but wasteful; it is dominated by discordance totals the observed accuracies rule out. We therefore restrict *v* to a (1 − *γ*) confidence region built by Bonferroni allocation from two-sided Clopper–Pearson intervals [45] for the marginals, mapped through the Fréchet cap, and add *γ* to the p-value, following Berger and Boos [44].

Two numerical steps could break the guarantee without any visible symptom. A grid maximum is a *lower* bound on a supremum, so a grid p-value can be too small. And a grid scan over Δ_0_ can step across a narrow accepted region, which matters because the acceptance set is not known to be connected. We remove both by working with rectangular cells in (Δ, *v*) and bounding the p-value over an entire cell. On [Δ_*L*_, Δ_*U*_] × [*v*_*L*_, *v*_*U*_], with *u* = *v* + Δ and *w* = 1 − 2*v* − Δ,

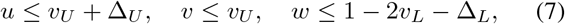

so every monomial *u*^*b*^*v*^*c*^*w*^*r*^ is dominated by the product of those endpoints and a termwise sum bounds each tail over the whole cell.

### Algorithm 1

DentalPair‑Cert. Inputs: *x*_1_, *x*_2_, common denominator N, level *α*, budget *γ*, nuisance mesh *hv*, tolerance *εΔ*. Output: an interval containing the exact Berger–Boos confidence set for Δ.

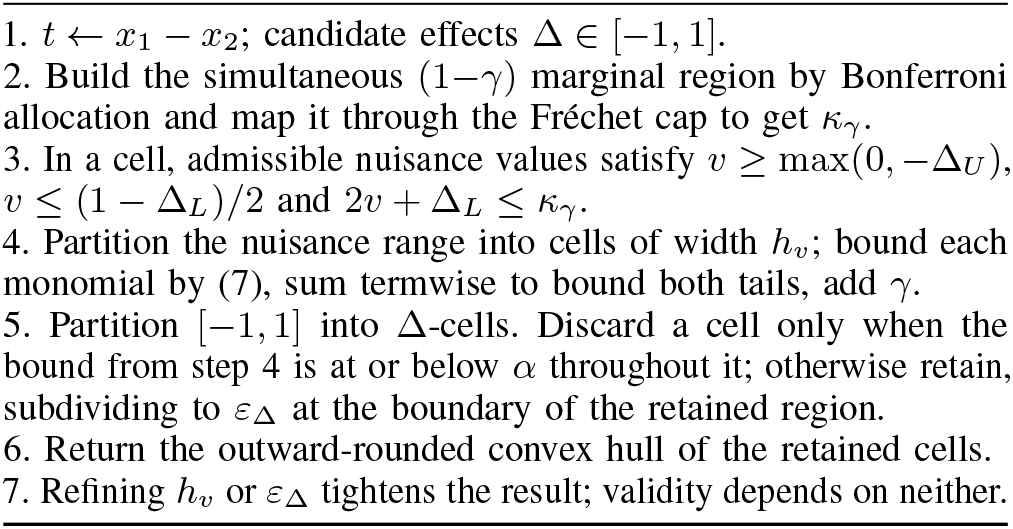

### Proposition 2

(Certified outer inversion). *If a* Δ*-cell is discarded only when a rigorous upper bound on the Berger– Boos p-value is at or below α for every* Δ *in that cell, then the union of retained cells contains the exact Berger–Boos confidence set, and its convex hull has coverage at least* 1 − *α*.

*Proof sketch*. Any Δ whose p-value exceeds *α* cannot lie in a cell whose valid uniform upper bound is at or below *α*, so it is never discarded. The returned set is an outer approximation of the exact acceptance set, and enlarging a set cannot reduce coverage. Refinement of *h*_*v*_ or *ε*_Δ_ changes tightness, not validity.

□

Two properties keep the computation affordable. The p-value depends on the data only through (*x*_1_, *x*_2_), and the cell bound depends on the parameters only through (*π*_10_, *π*_01_). Coarse meshes run first, and refinement is spent only on cells a coarse bound cannot already reject, and only at the boundary of the retained region, since interior cells cannot move the hull.

## VII. Experiments

### A. Three layers of evidence

The claims in this section rest on three different kinds of evidence, and conflating them would misrepresent all three. **(A)** The coverage guarantee for DentalPair-Cert is proved: Proposition 2 together with the certified outer bounds. **(B)** Exhaustive finite-sample coverage is *computed* at *N* ∈ {10, 15, 20, 25, 30}, where every compatible table can be weighted by its exact multinomial probability, so those figures carry no Monte Carlo error. **(C)** Operating characteristics over 4,200,000 replicates are *estimated* by simulation, for the methods that were actually run over that grid. DentalPair-Cert was not among them: the grid was executed with a grid-based, uncertified Berger–Boos implementation, and we report that procedure under its own name in Table IV rather than attributing its numbers to the certified algorithm. Nothing in Table IV is a theorem bound, and no cell in it is filled from a guarantee.

### B. Validity

The guarantee comes from Proposition 2 together with the certified numerical bounds, not from any experiment. What the experiments establish is that the implementation does what the proposition describes, and the strongest available check is exhaustive rather than simulated. For small *N*, coverage is a finite sum, so every table (*a, b, c, d*) summing to *N* can be weighted by its exact multinomial probability and the coverage computed rather than estimated.

We do this at *N* ∈ {10, 15, 20, 25, 30} under 7 true joint distributions spanning strong negative dependence, near independence, strong positive dependence, high accuracy in both arms, near-boundary probabilities, an exact null and a large true difference. The enumeration comprises 2,455 certified intervals, one for every observable (*x*_1_, *x*_2_). Minimum coverage is 97.49%, never below the nominal 95%; Table III gives the full breakdown. The worst case sits at the smallest sample size under strong negative dependence, which is where the variance gap (3) is widest, and coverage rises towards one as the dependence turns positive. That pattern is expected: the procedure is uniform over the dependence, so it pays most where the dependence is least favourable.

Seven implementation tests pass, covering the feasibility limits, the Clopper–Pearson and Bonferroni construction, normalisation of the tail mass at true probability triples, monotonicity of the certified bound under mesh refinement, a property test that the cell bound never falls below direct evaluation inside the cell, and a global inversion test against a dense reference computation.

**TABLE III.**
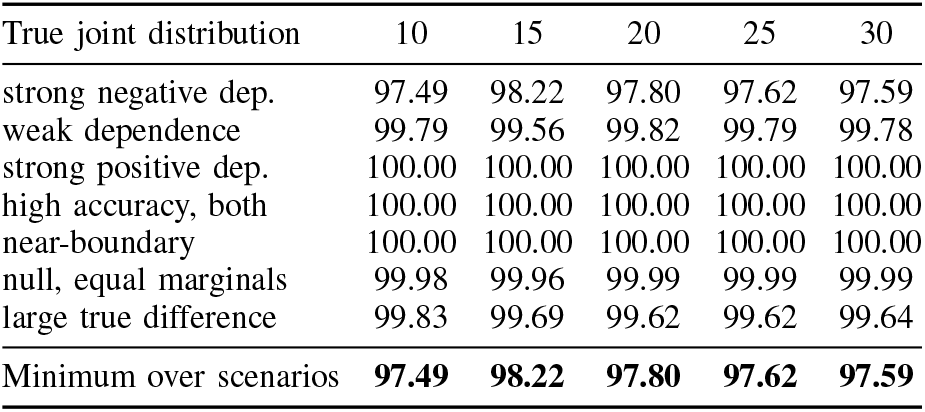
Exhaustive finite-sample coverage of DentalPair-Cert (%), evidence layer B. Every table (*a, b, c, d*) summing to *N* is weighted by its exact multinomial probability, so these are computed coverages carrying no Monte Carlo error. Columns are *N*; nominal level 95%.

| True joint distribution | 10 | 15 | 20 | 25 | 30 |
| --- | --- | --- | --- | --- | --- |
| strong negative dep. | 97.49 | 98.22 | 97.80 | 97.62 | 97.59 |
| weak dependence | 99.79 | 99.56 | 99.82 | 99.79 | 99.78 |
| strong positive dep. | 100.00 | 100.00 | 100.00 | 100.00 | 100.00 |
| high accuracy, both | 100.00 | 100.00 | 100.00 | 100.00 | 100.00 |
| near-boundary | 100.00 | 100.00 | 100.00 | 100.00 | 100.00 |
| null, equal marginals | 99.98 | 99.96 | 99.99 | 99.99 | 99.99 |
| large true difference | 99.83 | 99.69 | 99.62 | 99.62 | 99.64 |
| Minimum over scenarios | **97.49** | **98.22** | **97.80** | **97.62** | **97.59** |

**TABLE IV.** Observed operating characteristics over 210 conditions and 4,200,000 comparisons (evidence layer C). Every cell is a Monte Carlo minimum or maximum from methods actually run on the whole grid; none is a theorem bound. Coverage is the minimum over the grid, type-i error the maximum over true-null conditions. Methods above the rule need the paired table; those below need only the two accuracies and *N*. DentalPair-Cert is deliberately absent: it was not run on this grid, and its guarantee is layer a with exhaustive support in layer B (TABLE III).

| Method (all simulated) | Min. cov. (%) | Max. type-I (%) |
| --- | --- | --- |
| Paired Wald | 60.0 | — |
| Newcombe square-and-add | 92.6 | — |
| Tango score | 93.5 | 5.5 |
| Exact McNemar | — | 4.5 |
| Independence Wald | 74.5 | 12.2 |
| Maximum-variance Wald | 74.5 | — |
| Bonferroni–Clopper–Pearson | 99.3 | 0.7 |
| Grid Berger–Boos (uncertified) | 97.97 | 2.02 |

### C. Operating characteristics

Table IV reports a factorial simulation of 210 conditions with 20,000 replicates each, 4,200,000 comparisons in all, crossing seven accuracy regimes with five sample sizes and six dependence settings. Seven methods were evaluated on every replicate; every value in the table is an observed Monte Carlo minimum or maximum from that run. The independence analysis is the worst performer on both axes, at 74.5% minimum coverage and 12.2% maximum type-I error, with its worst case at mid-range accuracy under negative dependence and *N* = 25, which is where the variance gap (3) is largest. The maximum-variance plug-in interval, a natural first fallback, falls to 74.5%; it is a useful diagnostic but not a confidence procedure. Among methods needing the paired table the score interval is best calibrated at 93.5% coverage and 5.5% type-I error. The grid Berger–Boos procedure, which is the uncertified ancestor of the algorithm in Section VI, held 97.97% coverage on this grid; that is an observed value for that implementation and is not evidence about DentalPair-Cert, whose grid maximum is replaced by a certified outer bound precisely because a grid maximum can be too small.

### D. Cost of the certified interval

At *N* = 200 with *x*_1_ = 172 and *x*_2_ = 156, DentalPair-Cert returns [ −0.035, +0.193] of width 0.228, using *κ*_*γ*_ = 0.578 and 348 certified cell evaluations at *γ* = 0.001, *h*_*v*_ = 0.0005 and *ε*_Δ_ = 0.0005. The Bonferroni–Clopper–Pearson interval, the assumption-free comparator evaluated here, has width 0.250. The maximum-variance Wald interval is narrower at 0.165 but carries no finite-sample guarantee. Median wall-clock time was 14.3 s per interval at *N* = 200 on one core, with a 95th percentile of 41.5 s. Fig. 3 places these side by side. The independence interval is the narrowest of the four and the only one excluding zero, which is the whole difficulty in one picture: the method that is not valid here is also the one that returns the answer an author would prefer. A certified interval is wider than the paired interval the missing integer would have permitted. That difference is the price of the omission, and it is the argument for Theorem 1.

**Fig 3.**
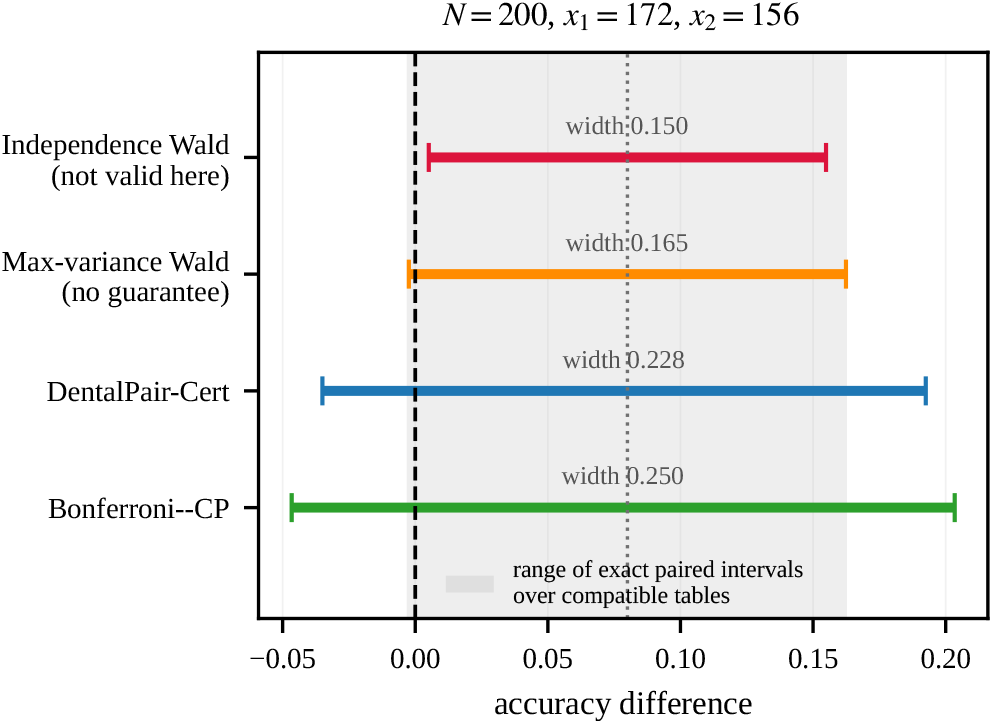
What each method returns on the same published pair. The shaded region spans the two-sided 95% Tango score interval, located by bisection over all compatible tables, so it is the range a reader could have been given had the joint table been printed. Widths are marked above each interval.

### E. Mesh refinement tightens without affecting validity

Section VI claims that *h*_*v*_ and *ε*_Δ_*S* are tightness and runtime parameters rather than validity parameters. Table V checks that on real inputs, at 2 sample sizes and 4 settings each. Two properties are asserted and both hold in every case. Each coarser setting returns an interval that *contains* the interval returned by every finer setting, so no choice of mesh can produce a result that is too short. And widths fall monotonically under refinement, from 0.3300 at *h*_*v*_ = 0.005 to 0.2275 at *h*_*v*_ = 0.0005 for the worked example. What refinement costs is runtime, which rises over the same ladder; the finest setting is the one used everywhere else in this paper, at a median 14.3 s per interval at *N* = 200 on one core (Python 3.11.15, NumPy 2.4.4). Making the procedure faster is engineering and forms no part of the validity argument.

**TABLE V.** Mesh refinement changes tightness, not validity. Each coarser setting returns an interval that strictly contains the finer ones, and widths decrease monotonically towards the finest computed reference. Enclosure and monotonicity verified in every case: yes.

| Case | $h_v$ | $\varepsilon_\Delta$ | lower | upper | width |
| --- | --- | --- | --- | --- | --- |
| $N = 200$ | 0.005 | 0.005 | -0.0850 | +0.2450 | 0.3300 |
|  | 0.002 | 0.002 | -0.0540 | +0.2120 | 0.2660 |
|  | 0.001 | 0.001 | -0.0420 | +0.1990 | 0.2410 |
|  | 0.0005 | 0.0005 | -0.0350 | +0.1925 | 0.2275 |
| $N = 282$ | 0.005 | 0.005 | -0.0950 | +0.1900 | 0.2850 |
|  | 0.002 | 0.002 | -0.0600 | +0.1580 | 0.2180 |
|  | 0.001 | 0.001 | -0.0480 | +0.1460 | 0.1940 |
|  | 0.0005 | 0.0005 | -0.0410 | +0.1390 | 0.1800 |

### F. Controlled paired benchmark on non-clinical image data

Everything above reasons about a table that reports withhold. This experiment produces one, on data where the full joint structure is available.

#### Data

The scikit-learn *Digits* dataset [18], [19]: 1,797 8 × 8 greyscale images of handwritten digits, ten classes, pixel intensities on a 0–16 scale. These are handwritten digits, not radiographs. The experiment is not evidence about dental diagnostic performance, about clinical effect sizes, or about any AI–dentist comparison. Its purpose is narrow and twofold: to show that models scored on shared cases have observable error dependence, and to quantify how much comparative information is lost when a *known* joint table is reduced to its marginals. Evidence that this dependence also holds at scale comes from published measurements on much larger shared test sets [40], [41], not from here.

#### Protocol

Ten classifier families and near-tied variants of the same family are trained on clean images and evaluated on corrupted ones, with predictions taken out-of-fold over a stratified five-fold split (seed 20260821), so every image is scored by every model exactly once and all models see identical test items. Corruption is additive Gaussian noise of SD 2.0 on the 0–16 intensity scale, applied once with the same seed; translation was tried and discarded because at 8 × 8 a one-pixel shift drives every model to near chance, which measures the corpus rather than model agreement. The noise level was fixed before any dependence or interval quantity was computed, chosen so the surviving models span a realistic accuracy band. Models falling below an accuracy floor of 0.4, also fixed in advance, are excluded as not being credible competitors; 10 arms remain, giving 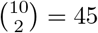 ordered-by-accuracy pairs, each contributing one 2 × 2 table.

Across 45 pairs the dependence is positive in 45/45 of them (Fig. 2, right), with *ϕ* from 0.099 to 0.856 and median 0.342. The rate at which both arms fail the same item exceeds its independence value by a median of 0.0111. Comparing what a reader could compute with the table against what the marginals alone support, the widest compatible interval exceeds the true paired interval by a median factor of 1.24 and by as much as 2.30.

In 3 pairs the marginals cannot settle the verdict. The sharpest case is a difference of 0.50 accuracy points at *ϕ* = 0.767. With the joint table the difference is significant, the paired interval having width 0.0108. Without it, 42 tables are compatible and the widest has width 0.0212 and contains zero. A reader given only the two accuracies and *N* could not have reached the conclusion the data support.

### G. Panels and clustered units

Feeding the measured joint-correct counts into the linear program of Section V gives the reductions reported there: 57% and 37% median reduction in the certified discordance cap for pairs with and without the reference, from 6 published integers.

The certified procedure needs an integer count out of the common denominator, so with several readers it applies to each fixed reader separately rather than to an averaged pseudo-count, which is not an integer count of anything. For 4 readers at *N* = 200 the certified widths had median 0.226 against 0.133 for the covariance-aware analysis available when reader-level data are published, a median ratio of 1.69. Inference about a reader *population* is a different target requiring reader sampling to be modelled, and remains a multi-reader multi-case problem [32].

Clustering is a separate layer, and dental data are clustered by construction: teeth sit inside patients. Under a shared-effect simulation with 40 patients and 8 teeth each, a patient effect loading unequally on the two arms drove paired Wald coverage to 79.3% when clustering was ignored; making the patient the sampling unit restored 94.5%. No procedure addressed to AI-dentist dependence within a unit can repair a misidentified sampling unit.

## VIII. The dental literature

### A. What recent studies make recoverable

We examined 9 recent comparative AI-versus-reader studies and recorded, for each, what the published report makes recoverable about the joint structure. This is a purposive sample of studies we located and read. It is not a systematic review, and it supports no estimate of how common any pattern is in the field; for population-level statements we rely on the published appraisals cited in Section II. Table VI lists them individually and Table VII gives the tally.

**TABLE VI.** What each of 9 recent comparative ai-versus-reader studies makes recoverable about the joint structure. “Recoverable” means a secondary analyst could attach comparative uncertainty to the ai-versus-reader difference. a purposive sample, not a systematic review.

| Study | Yr | Rdrs | What the report contains | Rec. |
| --- | --- | --- | --- | --- |
| [1] | 2025 | 6 | paired test on discordant units | yes |
| [8] | 2026 | 2 | covariance-aware head-to-head | yes |
| [5] | 2025 | 3 | AI–clinician agreement rate | yes |
| [4] | 2023 | 9 | paired AUC test, counts absent | no |
| [6] | 2024 | 2 | cross-tab against human reference | no |
| [3] | 2025 | 6 | per-arm accuracies only | no |
| [2] | 2025 | 60 | per-arm accuracies only | no |
| [7] | 2026 | 2 | per-arm accuracies only | no |
| [9] | 2025 | 4 | per-arm accuracies only | no |

**TABLE VII.** What 9 recent comparative studies make recoverable about the joint AI-by-reader structure. a purposive sample, not a systematic review.

| What the report contains | Studies |
| --- | --- |
| Paired test on discordant units [1] | 1 |
| Covariance-aware head-to-head estimate [8] | 1 |
| AI–clinician agreement rate [5] | 1 |
| Paired AUC test, counts not given [4] | 1 |
| Cross-tabulation against a human reference [6] | 1 |
| Per-arm accuracies only | 4 |
| **Comparative uncertainty recoverable** | **3 of 9** |

Three of the 9 report something from which comparative uncertainty can be recovered, and one of those reports the paired test directly. The remainder publish quantities from which the joint table cannot be reconstructed. The distinction we want to draw is not between careful and careless work. Several of these studies are methodologically strong in every other respect; what they omit is a convention, not a control.

### B. Implications for study design

Table I also reads backwards, as a design constraint. An investigator who expects a three-point difference and plans a study of two hundred units should know in advance that, reported in the conventional way, the result may well not settle the question: at that size the boundaries are 1.50 and 7.00 points, so a three-point difference lands in the pairing-dependent zone. Two responses are available. Either enlarge the study until *d*_high_ falls below the difference worth detecting, which for a three-point difference means roughly a thousand units, or report the extra integer and plan the analysis around the paired test, in which case the conventional power calculation for McNemar’s test applies and the required sample is far smaller. The second is almost always cheaper. This is the practical reason the recommendation is not merely a matter of bookkeeping: it changes what a feasible study can conclude.

### C. Reader composition moves the estimand

One further caution belongs with any AI-versus-dentist claim. Re-expressing the sensitivities reported by Zhu *et al*. [4] with the AI system and image set held fixed, the mean AI-minus-dentist difference was −0.041 against senior dentists, +0.010 against mid-career readers and +0.053 against junior readers. This is a descriptive pattern from a single study and generalises to nothing. It does show that who was recruited changes what “the AI beats the dentist” means, a question of estimand definition rather than of statistics.

The same three numbers also locate the decision zones in a real study rather than an abstract one. At that study’s size the boundaries are 1.06 and 6.03 accuracy points (Table I). The senior-reader difference of 4.11 points and the junior-reader difference of 5.32 points both fall in the pairing-dependent zone, so the published marginals settle neither; the mid-career difference of 0.97 points falls below the lower boundary and is non-significant under every compatible table. So 2 of 3 comparisons in one audited study sit where the reported numbers do not determine the answer. That is a fact about these three comparisons, not a frequency in the literature.

## IX. Discussion

### A. What to report

The recommendation is one integer per comparison, or *R* integers for a panel organised around a reference reader. A results table comparing an AI system with a dentist should carry one more column beside “AI: *x*_1_*/N*” and “dentist: *x*_2_*/N*”: the number of units both classified correctly. With it a reader computes McNemar’s test or a Tango interval directly [25], [27], [29]. Without that count the conventional paired variance and the standard paired interval are not identified, though dependence-robust inference such as DentalPair-Cert remains available. Releasing per-unit results is strictly better and subsumes the recommendation, but it is a heavier ask, and one integer captures most of what a reader needs for the comparison the paper is actually making.

### B. A worked example of the recommendation

It is worth seeing how little the change involves. A study of *N* = 282 units reporting the AI correct on 245 and the dentist on 231 leaves 38 compatible tables and an interval ranging from 0.0518 to 0.1309 in width. Adding one column to the results table, “correct in both arms: *a*”, fixes the table uniquely by (4); McNemar’s test and the Tango interval then follow directly. No extra images, no extra readers, no re-analysis: the number is already in the scoring script.

Table VIII sets out what each level of reporting permits.

**TABLE VIII.**
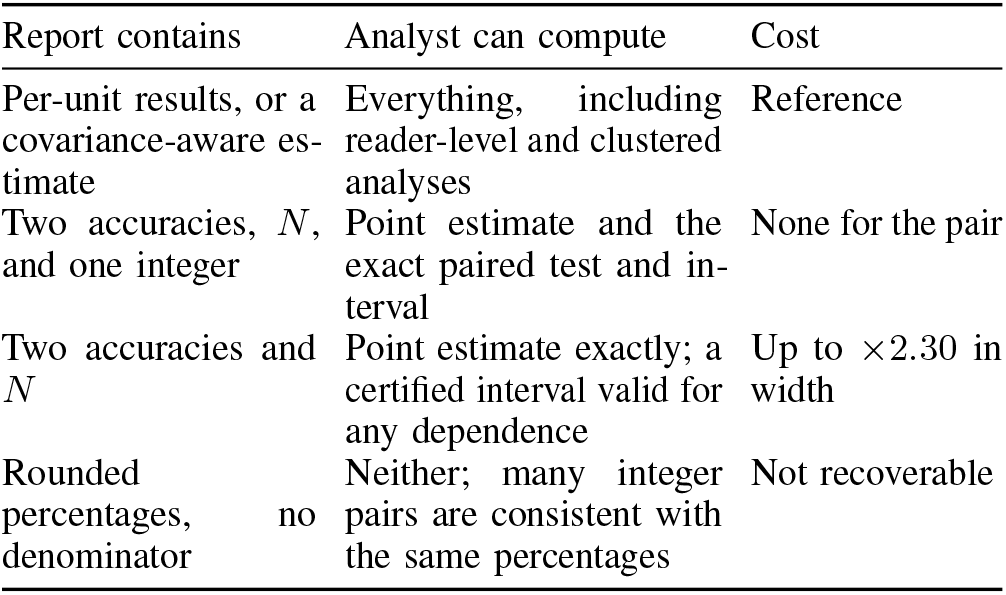
What each level of reporting permits a secondary analyst. The cost column is measured in this paper..

| Report contains | Analyst can compute | Cost |
| --- | --- | --- |
| Per-unit results, or a covariance-aware estimate | Everything, including reader-level and clustered analyses | Reference |
| Two accuracies, $N$, and one integer | Point estimate and the exact paired test and interval | None for the pair |
| Two accuracies and $N$ | Point estimate exactly; a certified interval valid for any dependence | Up to $\times 2.30$ in width |
| Rounded percentages, no denominator | Neither; many integer pairs are consistent with the same percentages | Not recoverable |

### C. Scope

The method needs both arms to have read the *same* units with a common integer denominator. If the arms read different case sets, if denominators differ, or if *N* is unreported, none of this applies. Reconstructing *N* from a rounded percentage is not a substitute: a sensitivity given to three decimals is consistent with many integer count pairs. The analysis treats correctness as a Bernoulli outcome per unit and conditions on the case set; it says nothing about between-study heterogeneity or about variability in model training.

### D. Consequences for pooling

The problem compounds when studies are combined. A meta-analysis of AI-versus-dentist accuracy needs a within-study variance for each comparison, and that is precisely the quantity the marginals leave unidentified. The options are to treat the two arms as independent, which Table IV shows is anti-conservative in the direction that matters, or to pool the arms separately, which answers a different question. Reviews of this literature have repeatedly abandoned or qualified pooling on grounds of heterogeneity [11], [13], [14]; part of what is recorded as heterogeneity is an identification failure that between-study modelling cannot fix. A certified interval per study gives a defensible input where the integer is missing, and the integer itself removes the difficulty where authors can supply it.

### E. For reviewers and editors

The change is small enough to be a review comment. When a manuscript compares an AI system against clinicians on shared images, asking for the number of units correct in both arms costs the authors one query against data they already hold, and converts a comparison whose conventional paired uncertainty is unidentified into one where it is not. Where a reporting checklist is in use [20]–[22], it fits as a single additional field.

### F. Limitations

The guarantee is uniform over within-unit dependence between the two arms and over nothing else. Clustered units are a separate layer, as our simulation shows, and dental data are almost always clustered. The reader-panel bound replaces each covariance by the pairwise Fréchet endpoint that increases the variance; this is valid termwise but is not claimed jointly sharp for more than two arms, and sharpness would need a separate joint-attainability argument. The linear program is exponential in the number of arms and we do not run it beyond a moderate panel. The controlled experiment uses a small non-clinical benchmark and establishes that the joint structure is real and positive, not how large it is at clinical scale. The audit in Section VIII is purposive and supports no prevalence claim.

## X. Conclusion

An AI system and a dentist reading the same radiographs are being compared in a paired design, whether or not the paper reports it that way. Reporting two accuracies separately removes the information needed to identify the conventional paired uncertainty, while leaving the point estimate intact and looking authoritative. At the sizes dental reader studies use, the published numbers resolve a difference only outside a band: on a 282-unit study, differences between 1.06 and 6.03 accuracy points cannot be adjudicated from what is printed. One additional integer removes the problem for a pair, and *R* integers substantially remove it for a panel. Where that integer was never published, DentalPair-Cert supplies an interval that holds whatever the unreported dependence, at a cost we measure rather than assert. The cheapest of these options is also the best one: report the number.

## Data Availability

All data produced in the present study are available upon reasonable request to the authors

## Acknowledgements and disclosure

Generative AI assistance was used in preparing this work. Anthropic Claude contributed to deriving and implementing the DentalPair-Cert algorithm and the linear-programming bounds, to the validation experiments, and to drafting the manuscript. OpenAI ChatGPT assisted with code review and source triage in earlier drafts. No model version is named, because the authors cannot independently verify which version served those sessions. Every numerical result is produced by the accompanying scripts rather than by either system, and each is re-derived from its stored output by the verification harness released with the code. The authors are responsible for the derivations, the outputs and the citations; this statement will be updated to record their verification once it has been completed.

